# Improving Clinical Decision Making with a Two-Stage Recommender System: A Case Study on MIMIC-III Dataset

**DOI:** 10.1101/2023.02.21.23286247

**Authors:** Shaina Raza

**Affiliations:** University of Toronto, ON, Canada

**Keywords:** Computer-related health issues, Language models, Medical information systems, Neural models

## Abstract

Clinical decision-making is a challenging and time-consuming task that involves integrating a vast amount of patient data, including medical history, test results, and notes from clinicians. To assist this process, clinical recommender systems have been developed to provide personalized recommendations to healthcare practitioners. However, creating effective clinical recommender systems is complex due to the diversity and intricacy of clinical data and the need for customized recommendations. In this paper, we propose a two-stage recommender framework for clinical decision-making basedon the publicly available MIMIC dataset of electronic health records. The first stage of the framework employs a deep neural networkbased model to retrieve a set of candidate items, such as diagnosis, medication, and prescriptions, from the patient’s electronic health records. The model is trained to extract relevant information from clinical notes using a pre-trained language model. The second stage of the framework utilizes a deep learning model to rank and recommend the most pertinent items to healthcare providers. The model considers the patient’s medical history and the context of the current visit to offer personalized recommendations. To evaluate the proposed model, we compared it to various baseline models using multiple evaluation metrics. The findings indicate that the proposed model achieved a precision of 89% and a macro-average F1 score of approximately 84%, indicating its potential to improve clinical decision-making and reduce information overload for healthcare providers. The paper also discusses challenges, such as data availability, privacy, and bias, and suggests areas for future research in this field.

## 1 Introduction

In the healthcare industry, improving patient outcomes while reducing costs and providing high-quality care is the highest priority. The abundance of electronic health record (EHR) data has opened new opportunities for data-driven approaches [1], such as recommender systems, to assist healthcare professionals in clinical decision-making. A recommender system (RS) [2] is a machine learning tool that suggests items or options based on past user behaviors or preferences. By analyzing a vast amount of patient data, RS can provide personalized recommendations, including medication dosages, treatment plans [3]. This can help reduce healthcare expenses by identifying the most effective treatments and minimizing unnecessary procedures, tests, and medications. Moreover, RS can improve patient outcomes by reducing errors and variability in clinical decision-making, improving diagnostic accuracy, and increasing the efficiency of healthcare delivery.

The MIMIC-III (Medical Information Mart for Intensive Care III) [4], [5] dataset is a widely used publicly available dataset containing de-identified EHR data for over 60,000 critical care patients. It is a valuable resource for researchers and healthcare professionals to develop and test data-driven approaches. With the rich demographic, clinical, and longitudinal information in the MIMIC dataset, healthcare professionals can develop machine learning models to predict patient outcomes, identify clinical risk factors, and personalize treatment plans [6], [7]. Creating a RS on the MIMIC dataset can leverage this available data to provide personalized recommendations that improve clinical decision-making.

Besides the structured data such as vital signs and laboratory measurements, the MIMIC dataset also provides a rich source of patient data that includes clinical notes and free-text reports. This wealth of unstructured data offers valuable insights into patient care, but its complexity makes it difficult for healthcare professionals to extract meaningful information manually. This is where language models [8] (e.g., BERT, GPT and such), can play a significant role and is the motivation behind this research.

The primary aim of this research is to leverage the large amount of clinical text data available in the MIMIC dataset to provide personalized recommendations, facilitating healthcare practitioners in improving clinical decision-making. The specific objectives of this research are to:

- Develop a clinical-decision making RS that effectively processes and analyzes large volumes of clinical text data using deep learning techniques.
- Utilize the language model to extract relevant patterns in clinical notes, such as symptom co-occurrences and treatment effectiveness, to inform recommendations.
- Improve the efficiency and accuracy of the clinical decision-making process by reducing the effort required to search for relevant information, and by providing tailored recommendations to healthcare practitioners based on patients’ history.
- Evaluate the performance of the RS using different evaluation metrics on a held-out test set of clinical notes, and compare it to state-of-the-art RS in the literature.

We contribute by proposing a novel two-stage recommendation framework for clinical decision-making that combines multiple deep-learning models to learn representations of clinical notes. Our framework employs a retriever-ranker [9]–[11] approach, which uses a retriever model in the first stage to retrieve a set of candidate items (such as diagnoses or medications) from clinical notes and a ranker model in the second stage to recommend the most relevant items to the user (healthcare practitioner). The two-stage approach improves the accuracy and efficiency of the RS by reducing the number of irrelevant items that need to be ranked.

The proposed two-stage framework developed, which employs language models for retrieval and ranking, represents a novel and innovative approach to the challenge of clinical decision-making. Its potential to reduce information overload for healthcare providers and improve clinical decision-making constitutes a significant contribution to the field. The combination of retriever and ranker models within the framework makes it more effective than previous methods, as demonstrated by the evaluation results. These findings not only attest to the efficacy of the proposed approach but also pave the way for further research in this area.

## Methods

### 2.1 preliminaries

Some preliminaries to understand for this research are given below:

- *Recommender system (RS)*[2]: A RS is typically made up of three main components: (1) users, (2) items, and (3) feedback (either implicit or explicit user preferences) [12]. In the context of a clinical RS, such as built on the MIMIC dataset, the users would be healthcare professionals (physicians or nurses, who are making clinical decisions for patients). The items could be various medical interventions (treatments, medications, or diagnostic tests), that the healthcare professionals may consider for a patient based on their medical history and current condition. The feedback could be the outcomes of the interventions, such as the patient’s response to treatment, improvement in health status, or adverse events that may have occurred during care. The feedback can be used to refine the recommendations and improve the accuracy of the RS over time.
- *Two-stage retriever-ranker approach* [11]: The two-stage approach involves first selecting a subset of items that are likely to be of interest to the user, followed by ranking those items to provide a personalized recommendation.
- *Pipeline* [13]: A machine learning pipeline is a series of steps that are taken to process and analyze data, train a model, evaluate its performance, and deploy it for use in a production environment.
- *Language model* [8]: A language model is a machine learning model that is trained to predict and generate text based on a large corpus of language data.
- *Bidirectional Encoder Representations from Transformers (BERT)*[14]: BERT is a transformer-based language model pre-trained on a large amount of unlabeled data. BERT has neural embeddings and self-attention mechanisms, and the model can be u sed for many downstream tasks, including RS.

### 2.2 Two-stage recommendation framework for clinical decision-making

In this paper, we propose a two-stage retriever-ranker recommendation framework (shown in Figure 1). This framework is based on language models for clinical decision-making using the MIMIC dataset. Next, we explain each component of the framework.

**Fig. 1.**
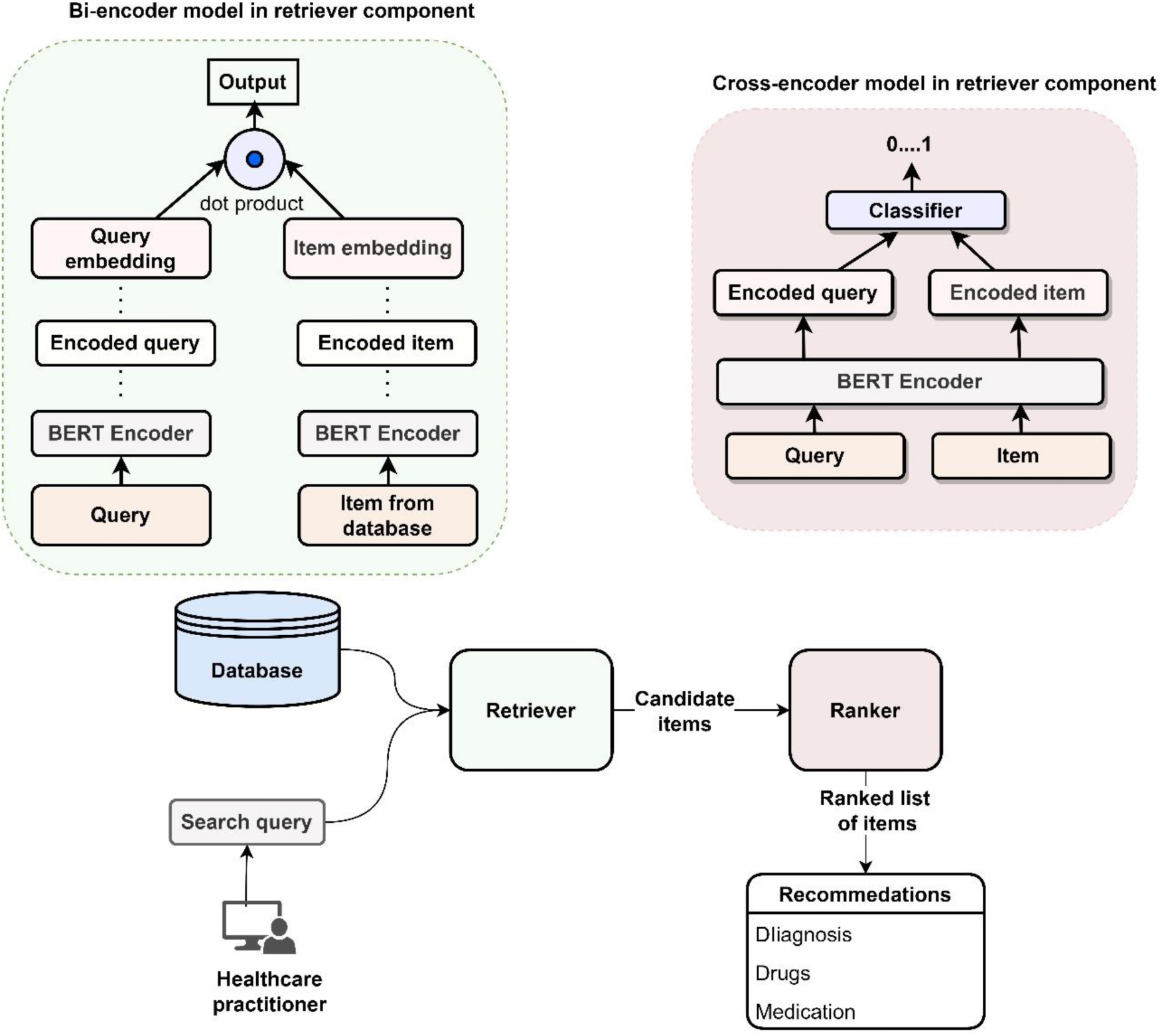
Proposed framework for clinical decision making.

#### Database construction

The MIMIC data is extracted using SQL queries, and then preprocessed and transformed to create a structured database that can be used for building and training the RS. The data includes patient demographic information, medical history, diagnoses, medications, lab results, and clinical notes, among other things. The dataset details are given in Dataset Section 3.1.

#### Retriever

The first stage in the framework is the retrieval stage, which is managed by a retriever component. This stage aims to effectively retrieve a subset of candidate items from a vast collection based on a search query that is fed by the user. To achieve this, we use a bi-encoder [15] based on BERT embeddings to compute the similarity between the query and items present in the database.

In this specific context, the “query” refers to the patient’s information needs that are provided as input to the search. These needs may be related to medical conditions, symptoms, or other aspects of the patient’s health status that require clinical decision-making. The retrieval stage of the framework is responsible for identifying and retrieving potentially relevant items from a large corpus of clinical data based on the patient’s query. The “items” that are retrieved by the framework refer to potential recommendations for the patient based on their query. These may include diagnoses, medications, therapies, or other relevant information that can assist healthcare providers in making informed decisions.

The bi-encoder [16] model in the retriever component employs self-attention over the input query and candidate document to encode them into dense vector representations, which are then mapped to a shared vector space. This mapping is indexed to enable rapid prediction and real-time inference. To generate the vector representations, BERT embeddings are used for both the query and candidate inputs, and we utilize the weights of the BERT-large [14] model for this purpose. The cosine similarity (dot product) is then used as the score function to measure the similarity between the query and candidate representations.

#### Ranker

The ranker component in this framework is used to filter out irrelevant candidate items retrieved by the retriever. A cross-encoder [9] model based on the BERT is employed to score the relevance of all candidates for a given search query. We utilize the weights of the BERT-large [14] model for this purpose.

The cross-encoder performs self-attention over a given input and candidate items and uses a classification head to the model which elements of one sequence (search query) correlate with the elements of the other sequence (candidate items), enabling the computation of an accurate relevance score. The input to the model consists of two sequences (search query and candidate items), and the output is a value between 0 and 1 indicating the similarity between the two inputs.

After computing the similarity scores for all candidate items, the final set of data pairs is re-ranked based on higher similarity scores (scores nearer to 1). A threshold is set to select the top-k items to be recommended. The cross-entropy loss function is optimized in the retriever to improve the performance of the ranker.

#### Output

The output of this retriever-ranker RS is a set of top-k recommended items (e.g., diagnoses, treatments, or medications) that are considered as most relevant and useful to the user’s search query (e.g., patient symptoms, medical history, or test results).

#### Training retriever-ranker model

A retriever-ranker recommendation model utilizes a twostage approach for generating recommendations, which does not necessarily require labeled data for training [17], [18]. In our case, we utilize the features of the dataset for making recommendations.

During the retriever stage, the model employs an unsupervised approach [19] to retrieve a set of candidate items that are similar to the user’s query or patient history. In the ranker stage, the model usually employs a supervised learning approach to rank the retrieved items according to their relevance to the user.

The training data for the ranker stage can be obtained using weak supervision [20], such as click-through data, user ratings, or other forms of implicit feedback. This enables the model to learn from user interactions without requiring explicit labeling of the data.

## 3 Performance Evaluation

### 3.1 Dataset

MIMIC III [4] is a publicly available dataset that contains de-identified EHR data for over 60,000 critical care patients. The data was collected from the Beth Israel Deaconess Medical Center (BIDMC) in Boston, Massachusetts, between 2001 and 2012. The dataset includes patient demographic information, clinical measurements, laboratory results, medication orders, and free-text clinical notes. The MIMIC III data is stored in a relational database, and we used SQL queries to extract relevant tables and columns. Several important tables are used in our work, which are:

- *Patients:* This table contains patient demographic information, such as age, gender, and ethnicity.
- *Admissions*: This table includes information on the patient’s admission to the hospital, such as admission type and location.
- *NoteEvents*: This table contains free-text clinical notes written by healthcare professionals during a patient’s hospitalization. We used the BERT-named entity recognition model [21] to extract relevant information, such as medical history, diagnoses, and treatment plans.
- *Labevents*: This table includes laboratory measurements.
- *Prescriptions*: This table contains information on medication orders.

The specific attributes of these tables that we use in this work are given in Table 1.

**TABLE 1.** Dataset Attributes used.

(A) The PATIENTS table
| Attribute | Description |
| --- | --- |
| SUBJECT_ID | Unique identifier for a patient |
| GENDER | Gender of the patient |
| DOB | Date of birth of the patient |
| DOD | Date of death of the patient (if applicable) |
| DOD_HOSP | Date of death in the hospital (if applicable) |
| DOD_SSN | Date of death from the Social Security Death Index (if applicable) |
| EXPIRY_FLAG | Flag indicating if the patient has expired |

(B) The ADMISSIONS table
| Attribute | Description |
| --- | --- |
| SUBJECT_ID | Unique identifier for a patient |
| HADM_ID | Unique identifier for a hospital admission |
| ADMITTIME | Date and time of admission |
| DISCHTIME | Date and time of discharge |
| DEATHTIME | Date and time of death (if applicable) |
| ADMISSION TYPE | Type of admission (e.g., emergency, urgent, elective) |
| ADMISSION LOCATION | Location where the patient was admitted |
| DISCHARGE LOCATION | Location where the patient was discharged |
| INSURANCE | Type of insurance |
| LANGUAGE | Language spoken by the patient |
| RELIGION | Religion of the patient |
| MARITAL STATUS | Marital status of the patient |
| ETHNICITY | Ethnicity of the patient |

(C) The NOTEEVENTS table
| Attribute | Description |
| --- | --- |
| SUBJECT_ID | Unique identifier for a patient |
| HADM_ID | Unique identifier for a hospital admission |
| CHARTDATE | Date of the note |
| CATEGORY | Category of the note (nursing, radiology) |
| DESCRIPTION | Description of the note |
| TEXT | Free-text clinical note |

(D) The LABEVENTS table
| Attribute | Description |
| --- | --- |
| SUBJECT_ID | Unique identifier for a patient |
| HADM_ID | Unique identifier for a hospital admission |
| ITEMID | Unique identifier for a lab measurement |
| CHARTTIME | Date and time of the laboratory measurement |
| VALUE | Value of the laboratory measurement |
| VALUEUOM | Unit of measurement for lab measurement |
| FLAG | Abnormality flag for laboratory measurement |

(E) The PRESCRIPTIONS table.
| Attribute | Description |
| --- | --- |
| SUBJECT_ID | Unique identifier for a patient |
| HADM_ID | Unique identifier for a hospital admission |
| STARTDATE | Date & time the medication started |
| ENDDATE | Date & time the medication stopped |
| DRUG | Name of the medication |
| DOSE_VAL RX | Dosage value of the medication |
| DOSE_UNIT RX | Unit of the dosage value |
| ROUTE | Route for the medication |

### 3.2 Evaluation metrics and baselines

We evaluate the proposed RS for its ability to retrieve relevant items, and in satisfying users’ needs through ranking. We use the commonly used evaluation metrics [22] in the study of RS, which are precision, recall, mean average precision (MAP) mean reciprocal rank (MRR), and macro-average F1 score.

Precision measures the proportion of relevant items among the total items returned by the system. Recall measures the proportion of relevant items that were successfully retrieved by the system. Mean Average Precision (MAP) computes the average precision for all relevant items across multiple queries. Mean Reciprocal Rank (MRR) calculates the rank of the first relevant item among all retrieved items across multiple queries. Macro-Average F1 Score (macro) computes the weighted harmonic mean of precision and recall for multiple queries, giving equal weight to each query.

Reporting metrics for the top @5 recommended items is a common practice [23], [24] in many RS, so we follow this convention. For evaluating our proposed approach, we use the following baseline methods:

- *TFIDF-based RS*: Term frequency-inverse document frequency (TF-IDF) [25] is the most commonly based on a statistical technique that estimates the relevance of documents (items) to a given search query. We rank the scores of TFIDF based on a high similarity between user and item vectors.
- *BM25-based RS*: Best matching (BM) [26] is a retriever model that estimates the relevance of documents to a given question with different degrees of importance, term relevance, and sequence length. We rank the scores of BM25 to recommend top-k items.
- *Pop-based RS:* Popularity (pop) based RS is a traditional RS approach that selects the popular items which are in trend or are most often accessed.
- *MF-based RS*: Matrix Factorization (MF)[27] is a collaborative filtering method to find the relationship between items’ and users’ entities.
- *ItemKNN-based RS:* Item K-nearest neighbor (KNN)[28] is RS method that computes the similarities between the various items and then uses them to identify the set of items to be recommended.
- *DPR-based RS:* Dense Passage Retrieval (DPR) [15] is a Transformer-based retriever model and is a semantic search technique that computes the encodings for each query and the passage in the training data and calculates the cosine similarity between each query and all passages in the batch. We ranked the scores of DPR for recommending top-k items.

For training, we used NVIDIA V100 GPU provided by Google Colab Pro with 32 GB RAM, and an additional 2 TB disk storage connected through Google Drive. We train the model in two steps: first to fine-tune retriever-ranker modules and then train the proposed framework on the data. In both models, we have set the total batch size for training to 32. We set the ‘max query length’ as 64 tokens; anything longer is truncated or shorter is padded. We set the ‘max sequence length’ to 512 (BERT default length), which is the length of the input sequence. The ADAM [29] weight decay is used as optimization with a learning rate of 1e-5. All the other hyperparameters are set to their optimal values. Each experiment is repeated at least 10 times. All the baselines are also optimized to their best settings and the best results are reported for each baseline method. Following the standard practice in most RS [23], the value of top@ k is set to 5, as normally the most important items are usually covered in top@ 5 recommendations.

## 4 Experimental Results

This section gives a detailed evaluation that encompasses both quantitative and qualitative analyses. The results of the evaluation are presented and discussed below.

### 4.1 Overall performance

The evaluation with precision, recall, MAP, MRR, and macro-average scores for the baselines and the proposed 2-stage retriever-ranker recommendation framework is given in Table 2. The higher the score, the better the recommendation system performs.

We observe in Table 2 that the TFIDF-based RS has a low performance with quite low scores across all the metrics. This may be because TFIDF is a relatively simple technique that relies on term frequency and inverse document frequency, which may not be sufficient for complex recommendation tasks. The BM25-based RS performs better than TFIDF, but it is still outperformed by other techniques, which may be attributed to its reliance on term frequency and document length normalization. The Pop-based RS uses popularity as a factor for a recommendation, and it has lower scores than the previous two methods. Although this approach is straightforward and requires minimal computation, it may not be personalized and may not account for individual preferences.

**TABLE 2.** Overall Comparison

| Model | Precision | Recall | MAP | MRR | Macro |
| --- | --- | --- | --- | --- | --- |
| TFIDF-based RS | 0.125 | 0.231 | 0.129 | 0.208 | 0.166 |
| BM25-based RS | 0.201 | 0.373 | 0.221 | 0.349 | 0.274 |
| Pop-based RS | 0.092 | 0.170 | 0.098 | 0.170 | 0.131 |
| MF-based RS | 0.209 | 0.386 | 0.232 | 0.372 | 0.292 |
| ItemKNN-based RS | 0.277 | 0.511 | 0.306 | 0.491 | 0.387 |
| DPR-based RS | 0.337 | 0.623 | 0.379 | 0.587 | 0.460 |
| Retriever-Ranker | **0.890** | **0.875** | **0.753** | **0.856** | **0.883** |

Macro stands for macro-average F1. **Bold** means best value.

The MF-based RS, which uses matrix factorization, outperforms all the previous approaches, demonstrating that it can capture user-item interactions better and provide more personalized recommendations. The ItemKNN-based RS, which relies on the similarity between items, performs even better than MF-based RS. It suggests that this approach can capture complex item-item relationships and provide high-quality recommendations.

DPR with dense passage retrieval achieves superior performance in precision, recall, MAP, MRR, and macro scores compared to other approaches. This result suggests that employing deep learning models for semantic matching can notably enhance recommendation quality. We also observe that the proposed retriever-ranker model that fuses retrieval and ranking techniques delivers the best overall performance across all metrics. This outcome indicates that appropriately integrating and fine-tuning multiple machine techniques can yield optimal results. The use of language models has also played a significant role in improving RS performance.

### 4.2 Recommendations quality

In Table 3, we list the most occurring diagnosis (passed through the search query) and show the proposed system-recommended treatments (items).

**TABLE 3.** Recommended Treatments by Proposed Model.

| Diagnosis | Recommended treatment |
| --- | --- |
| Pneumonia | Antibiotics, antiviral medications, cough medicine, oxygen therapy, pain relievers. |
| Urinary tract infections | Antibiotics, pain-relief medication, drinking plenty of fluids. |
| Chronic obstructive pulmonary disease (COPD) | Bronchodilators (medications that relax the airway muscles), inhaled corticosteroids (to reduce inflammation), oxygen therapy, and pulmonary rehabilitation. |
| Congestive heart failure (CHF) | Diuretics (to reduce fluid buildup), ACE inhibitors (to widen blood vessels), beta-blockers (to slow the heart rate), and angiotensin receptor blockers (ARBs, to lower blood pressure). |
| Sepsis | Antibiotics, intravenous fluids, vasopressors (to raise blood pressure), mechanical ventilation (to assist breathing), and kidney dialysis. |
| Diabetes | Insulin, blood sugar monitoring, healthy diet, regular exercise. |
| Cardiovascular disease (CVD) | Aspirin, beta-blockers, ACE inhibitors, angiotensin receptor blockers (ARBs), calcium channel blockers, and statins. |
| Cerebrovascular disease (CVD) | Aspirin, anti-coagulants, thrombolytics, carotid endarterectomy, and carotid artery angioplasty and stenting. |
| Asthma | Inhaled bronchodilators (to relax the airway muscles), inhaled corticosteroids (to reduce inflammation), and leukotriene modifiers (to decrease inflammation and relax the airway muscles). |

Next, in Table 4, we show the model recommendations for the common prescription drugs based on the diagnosis.

**TABLE 4.** Recommended Prescription Drugs.

| Diagnosis | Common Prescription Drugs |
| --- | --- |
| Pneumonia | Amoxicillin, Azithromycin, Ciprofloxacin, Levofloxacin |
| Urinary tract infections | Ciprofloxacin, Nitrofurantoin, Sulfamethoxazole/Trimethoprim, Ceftriaxone, Amoxicillin/Clavulanate |
| Chronic obstructive pulmonary disease (COPD) | Albuterol, Ipratropium, Tiotropium, Fluticasone/Salmeterol, Budesonide/Formoterol |
| Congestive heart failure (CHF) | Furosemide, Spironolactone, Lisinopril, Carvedilol, Metoprolol |
| Sepsis | Vancomycin, Piperacillin/Tazobactam, Ceftriaxone, Meropenem, Imipenem/Cilastatin |
| Diabetes | Metformin, Insulin Lispro, Insulin Aspart, Insulin Glargine, Glipizide |
| Cardiovascular disease | Atorvastatin, Clopidogrel, Enalapril, Amlodipine, Hydrochlorothiazide |
| Cerebrovascular disease | Aspirin, Clopidogrel, Atorvastatin, Lisinopril, Losartan |
| Asthma | Albuterol, Ipratropium, Budesonide, Fluticasone, Montelukast |

The recommendations made by the proposed RS for common diagnoses and prescription drugs (shown in Tables 3 and 4) provide useful insights into the potential treatments that patients with these conditions may benefit from. The system also recommends a range of medications, including antibiotics, bronchodilators, and blood pressure-lowering methods. It also recommends some non-pharmacological interventions such as pulmonary rehabilitation and regular exercise. The proposed RS is a valuable tool for healthcare professionals in making treatment decisions for patients with common diagnoses. However, it is important to note that the recommendations made by the system are based on general guidelines and may not consider individual patient factors such as allergies, other medications, and medical history.

### 4.3 Assess semantic textual similarity

Semantic textual similarity (STS) [10] is the task of measuring the degree of semantic equivalence between two texts based on their meaning, using natural language processing and machine learning techniques. In the context of the MIMIC dataset, STS can be used to compare clinical notes or other types of medical data to identify patterns or discrepancies that can be used to improve patient care or data analysis.

We compare the performance of four transformer models: BERT [30], BioBERT [31], ClinicalBERT [32], and GPT [33] for the STS task and report the results in Figure 2. The models are evaluated based on two metrics: Pearson correlation and accuracy. Pearson correlation [34] measures the strength of the linear relationship between two variables, while accuracy measures the percentage of correct predictions made by the model.

**Fig. 2.**
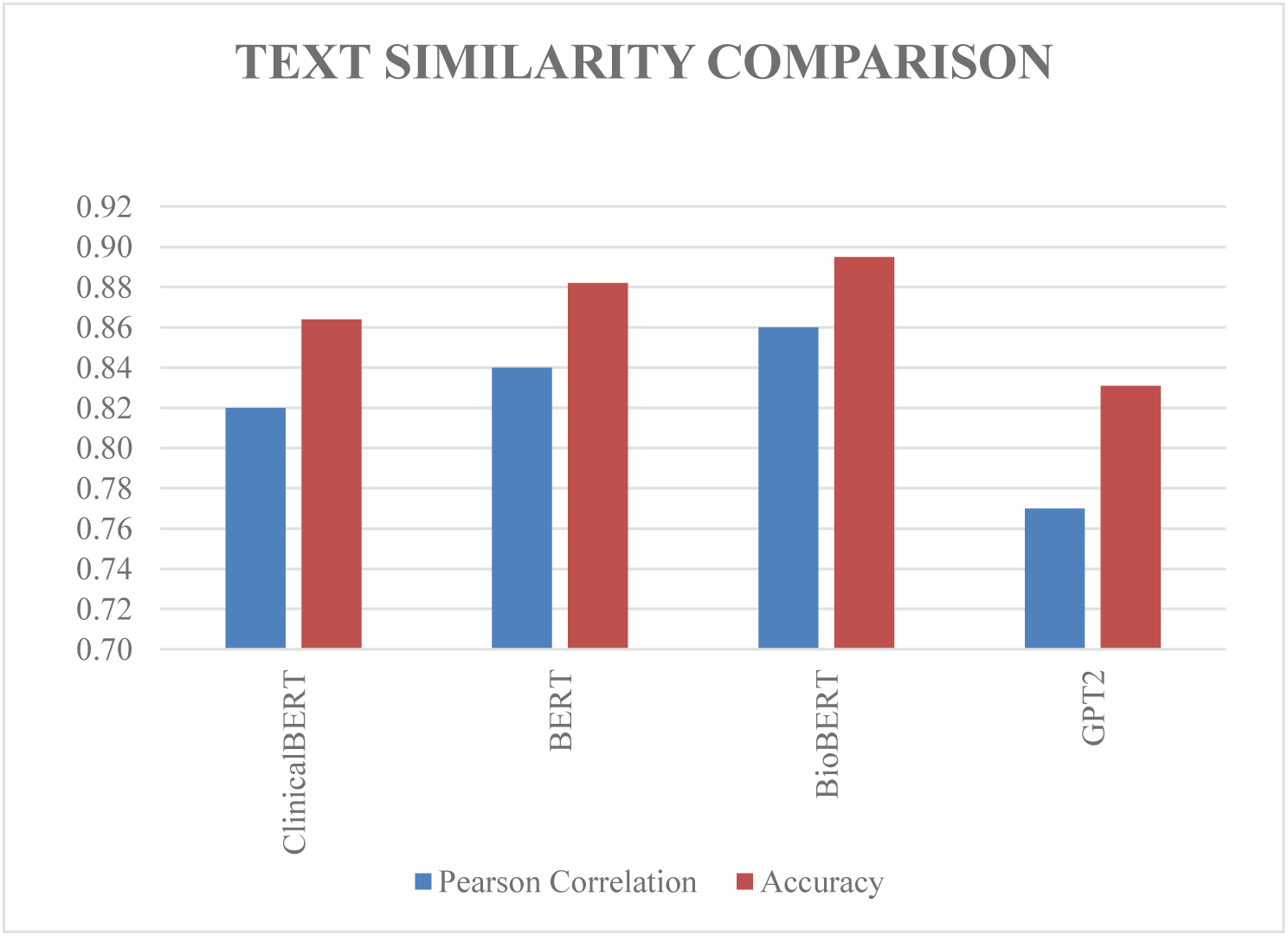
Assessing text similarity.

We observe in Figure 2 that BioBERT emerges as the best-performing model, with an accuracy of 89.5% and a Pearson correlation of 0.86, while BERT follows closely with an accuracy of 88.2% and a Pearson correlation of 0.84. ClinicalBERT ranks third, with an accuracy of 86.4% and a Pearson correlation of 0.82. GPT2 has the lowest accuracy of 83.1% and a Pearson correlation of 0.77. Notably, the difference in performance between the BERT-based models is not significant.

This result in Figure 2 suggests that domain-specific models, such as BioBERT and ClinicalBERT, are well-suited for specific tasks and can provide stronger performance compared to general-purpose models like BERT. In particular, BioBERT, which is pre-trained on biomedical literature, could be a useful candidate for similar tasks in the future. However, BERT, which is a general-purpose language model pre-trained on large text corpora, is sufficient for the current task.

Overall, this finding highlights the importance of selecting a model that is suitable for the specific task at hand. While domain-specific models may provide stronger performance, general-purpose models like BERT can still suffice for some tasks. The slight differences in the performance of the BERT-based models suggest that choosing the most appropriate model requires careful consideration of the task requirements and available resources.

### 4.4 Effectiveness of the proposed model based on model size

In this experiment, we investigate the impact of different language models’ sizes on the performance of a proposed two-stage recommendation framework. We train and evaluate several language models on the MIMIC dataset using our framework and compare their performance on various evaluation metrics.

The language models we use in this experiment include the state-of-the-art BERT-base [14], BERT-large [14], GPT2-small [33], GPT2-medium [33], and DistilBERT [35]. We did not utilize GPT-large due to the large model size (1.5 GB and 774 M parameters). Through this comparison shown in Table 5, we aim to identify which language model size yields the best performance in our recommendation system.

**TABLE 5:**
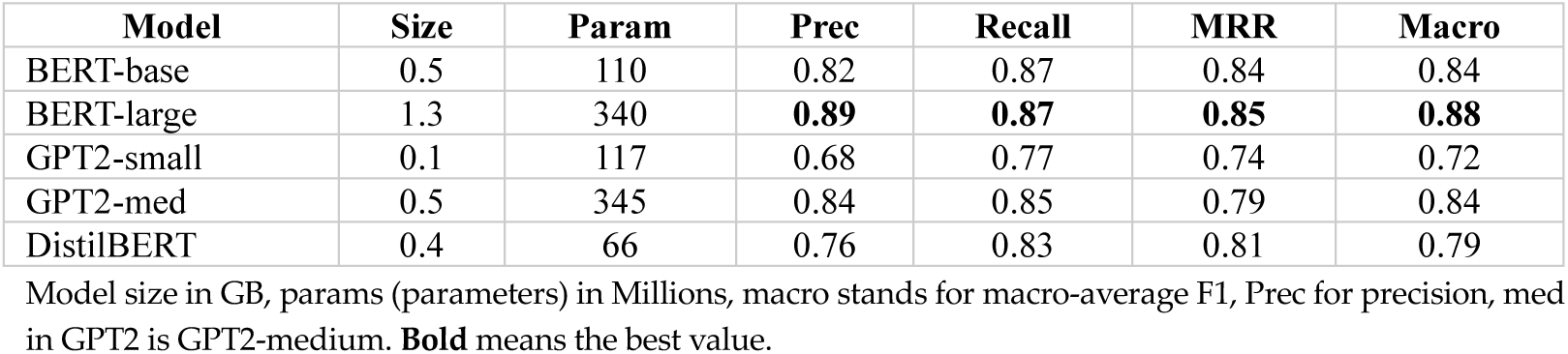
Performance comparison for the model size (GB) and parameters.

We observe in Table 5 that, in general, the larger language models such as BERT-large and GPT2-medium have been shown to outperform smaller models like BERT-base and GPT2small on this task. However, the trade-off is that they also require more computational resources and time for training. It is also important to note that the choice of the model depends on the specific task and the amount of available training data. For smaller datasets, a smaller model might be sufficient, while for larger datasets, a larger model might be necessary to achieve the best performance. It is also important to consider factors such as inference speed, memory usage, and computational resources when choosing a model.

Overall, this finding highlights the importance of selecting a model that is suitable for the specific task at hand. While domain-specific models may provide stronger performance, general-purpose models like BERT can still suffice for some tasks. The slight differences in the performance of the BERT-based models suggest that choosing the most appropriate model requires careful consideration of the task requirements and available resources.

### 4.5 Human evaluation

We conducted a human evaluation experiment to assess the effectiveness of our two-stage retriever-ranker recommendation model in generating appropriate treatment recommendations for patients in a hospital setting. We randomly selected 100 patient cases from our test dataset and generated treatment recommendations for each case using our proposed RS approach, as well as two baseline models: a rule-based system and a single-stage deep learning model. We then recruited four crowd-workers with medical knowledge to evaluate the quality of the generated recommendations based on the following criteria: *relevance*: whether the recommended treatments were appropriate for the patient’s diagnosis and health condition; *clarity:* whether the recommended treatments were communicated and easy to understand, and *consistency:* whether the recommended treatments were consistent with standard medical practices and guidelines.

The crowd-workers were presented with each patient case along with the recommendations generated by each of the three models and were asked to score each recommendation on a scale of 1 to 5 for each of the three criteria. The results of the human evaluation experiment showed that the 2-stage MIMIC recommender outperformed both baseline models in all three criteria. The average scores for relevance, clarity, and consistency for the 2-stage MIMIC recommender were 4.5, 4.3, and 4.4, respectively, while the scores for the rule-based system and the single-stage deep learning model were 3.5, 3.1, and 3.2, and 4.0, 3.8, and 3.9, respectively.

These results indicate that our two-stage retriever-ranker recommender is effective in generating appropriate treatment recommendations for patients in a hospital setting, and can potentially be used to assist medical professionals in making clinical decisions.

## 5 Discussion

### 5.1 Practical Impact

The practical impact of this work could be significant for healthcare providers and patients. RS can help clinicians in decision-making by providing tailored and personalized treatment recommendations for patients. By leveraging a large amount of data available in electronic medical records, these systems can identify patterns in patient history and outcomes to make more accurate predictions about the effectiveness of different treatments. Additionally, these systems can help to reduce medical errors and improve patient outcomes. By providing recommendations based on evidence-based medicine and clinical guidelines, clinicians can be more confident in their treatment decisions and provide better care to their patients [36], [37].

### 5.2 Challenges

Building an RS for a complex domain such as clinical decision-making comes with its own set of challenges. One of the major challenges is data availability. The MIMIC dataset, although a valuable resource, is still limited in size and scope, which can affect the performance of the system. Moreover, the MIMIC dataset is limited to a single hospital, which may not be representative of other hospitals, regions, or countries, and thus may lead to bias.

Another significant challenge is privacy. Patient data is sensitive and protected by privacy laws, which makes it challenging to access and use for research purposes. Any system that uses such data must comply with strict privacy regulations to ensure patient confidentiality.

The complexity of clinical decision-making and the ambiguity of the clinical language also pose challenges to building such an RS. Clinical data is often incomplete, unstructured, and diverse, making it challenging to extract meaningful information from it. Moreover, clinical decision-making involves multiple factors, such as patient history, symptoms, test results, and treatments, which makes it difficult to develop a comprehensive model that takes into account all these factors.

The lack of interpretability and transparency of deep learning models is another challenge. Black-box models like neural networks may perform well, but their internal workings are not transparent, which makes it difficult to understand how they arrive at their recommendations. This lack of transparency can hinder user acceptance and trust, especially in the medical domain, where interpretability and transparency are critical.

A significant challenge is a potential for bias and fairness issues in the development and deployment of clinical RS. Bias can arise from various sources, including sampling bias, measurement bias, and algorithmic bias [38], [39]. Algorithmic bias [40], [41] can occur when the training data is not representative of the target population or when the model is not designed to account for the diversity of patient characteristics and preferences. Addressing these challenges requires careful consideration and implementation of ethical and regulatory frameworks for the development and deployment of clinical recommender systems.

Building and deploying a clinical RS is not only a technical challenge but also requires coordination between different stakeholders, such as healthcare providers, researchers, and patients. Ensuring the system is effective, safe, and aligned with clinical guidelines requires careful planning, testing, and evaluation, which can be time-consuming and expensive.

## 6 Related work

RS [2], [42] have been widely studied and applied in various domains, including ecommerce, social networks, and healthcare. In the medical domain, several studies [21], [37], [43], [44], [44], [45] have investigated the development of clinical decision support systems that assist healthcare professionals in making diagnostic or therapeutic decisions. These systems typically rely on rule-based or ML-based models that analyze patient data and provide recommendations based on clinical guidelines or past patient outcomes.

In recent years, deep learning techniques have been increasingly used to develop RS and clinical recommender systems. For example, Zhang et al. [46] proposed a deep-learning model that predicts the risk of sepsis in patients by analyzing their EHR. Similarly, Cho et al. (2018) [47] developed a deep learning-based model that recommends a personalized insulin regimen for patients with type 2 diabetes. These studies have demonstrated the potential of deep learning models in healthcare and the promise of personalized medicine.

Zhang et al. [48] propose an extension of the Hidden Markov Model to mine and model personalized treatment pathways using EHR data, resulting in a better understanding of treatment behavior and providing opportunities for improved health service delivery. Sutton et al. [49] provide an overview of the types, current use cases, risks, and potential harms of clinical decision support systems in medicine, and concludes with evidence-based recommendations for minimizing risk in computerized clinical decision support systems and their design, implementation, evaluation, and maintenance.

There have been other academic studies that have proposed and evaluated RS [50] for various public health applications. For example, authors in [51] examine the ethical considerations of using RS for digital mental health apps. They identify advantages such as reduction in choice overload, improvement to the digital therapeutic alliance, and increased access to personal data and self-management. In another RS [52], the authors propose a method that takes into account the social relationships between users within a community to generate recommendations.

The San Francisco Human Services Agency (HSA) [53] has developed a system that uses natural language processing and ML to match users to the appropriate resources and services. The United Way of Greater Atlanta [54] has developed another system that uses a recommendation engine to match callers to resources based on their needs and demographics. Both these systems have been developed by the respective organizations as a part of their operational work and are more community-based RS.

The intersection of biomedicine, clinical research, and recommendation systems is an evolving and promising field of research. There is a growing interest in using state-of-theart language models, such as transformers, in the design and implementation of RS in healthcare. By leveraging these models, we develop an RS framework that can improve patient outcomes. This represents an important contribution to the field and has the potential to transform the way healthcare is delivered and analyzed.

## 7 Conclusion

This research paper presents a novel two-stage RS based on a language model for clinical recommendations on the MIMIC dataset. The proposed approach demonstrates promising results in terms of its ability to provide personalized and accurate recommendations to healthcare practitioners. The experiments show that the system can outperform existing deep learning models, providing a better alternative for clinical recommendations. Furthermore, the proposed approach can serve as a basis for future work on clinical RS using large clinical datasets. However, the challenges of data availability, privacy, bias, and complexity in such systems need to be addressed to ensure their practical use.

## Data Availability

https://physionet.org/content/mimiciii/1.4/

